# Philosophical Studies of Non-Pharmacological Pain Management with Transcultural Nursing Approach

**DOI:** 10.1101/2022.07.01.22277133

**Authors:** Yunani, Moses Glorino Rumambo Pandin

## Abstract

This article will discuss the philosophical study of non-pharmacological pain management by looking at perspectives in ontology, epistemology and axiology. Pain is an unpleasant sensory and emotional experience associated with tissue damage. The impact of pain can generate tension that can stimulate the central nerve to release catecholamines that cause arterial and tachycardial constriction. This can increase the afterload and decrease *the cardiac output*. Pain is influenced by biological, psychological, and social factors at varying degrees. Pain is a subjective experience of the patient so in the provision of nursing care to pain patients, it is necessary to pay attention to the factors that affect pain among other cultures. Understanding and knowledge of transcultural nursing theory, one of which is the Theory of Diversity-Based Diversity and Cultural Universality by Madeleine M. Leinenger with the sunrise model, must be owned by the nurse in conducting the assessment, making the diagnosis and arrange nursing intervention. Non-pharmacological pain management also needs to be done by nurses in overcoming pain problems in patients.

## 1. INTRODUCTION

Pain is something very complex, so the nurse must consider all the factors that influence the patient in feeling pain. This is very important as an effort to ensure that nurses use a holistic approach in the assessment and treatment of patients who experience pain. Some sources of stimulus that can cause pain include infections, cancer, diabetes, autoimmune diseases, suppression of nerve roots in the spine and several other diseases.

The impact of pain can generate tension, which can stimulate the central nerve to release catecholamines that cause arterial and tachycardial constriction. This can increase afterload and decrease *cardiac output* (Smeltzer et al., 2010). Pain can also result in the patient avoiding actions to prevent mobilization and performing deep breath exercises. The condition causes the patient to be unable to perform effective breathing, thereby reducing the ability to inspire and the limited effective cough that causes the airway to be ineffective. A poor pain control delay can affect hemodynamics which can trigger the occurrence of myocardial ischemia and other complications. The incidence of severe pain can extend the treatment period and hinder recovery. Improper pain care will have an impact on all human dimensions and impaired physical activity.

Race, culture and ethnicity are important factors in a person responding to pain. Such factors affect all sensory responses, including responses to pain. Problems can arise due to a person’s view of a member of the health team. Patients from certain cultural groups may have difficulty communicating to doctors and nurses because they come from different cultural backgrounds and ethnic groups. Health care providers may also have difficulty in assessing pain experiences from unknown cultural groups. Patients from different cultures may each be able to deal with pain in a variety of ways. However, it arises when the nurse is unable to recognize the way the patient responds to pain or when the nurse is unable to accept it. The study found that nurses’ assessments of patients’ pain experiences, were influenced by their belief values and culture. They also misinterpreted the patient’s pain experiences related to the primary language. Healthcare providers should be sensitive to supportive cultural factors and patient language in facilitating adequate pain management (Black & Hawk, 2014).

Efforts to overcome pain problems in patients several interventions in non-pharmacological pain management include: massage, hot and cold compresses, transcutaneous electrical nerve stimulation, acupuncture, acupuncture, deep breath, progressive relaxation, music, biofeedback and distraction (Black & Hawk, 2014). However, it is necessary to pay attention to several cultural factors in providing these interventions.

## 2. METHOD

The study used an integrative literature review method. This review proposed non-pharmacological pain management with transcultural nursing approach. We systematically searched Scopus, PubMed, ProQuest, and Science Direct. Search was using various combinations of keywords with the help of Boolean operators, including: “non pharmachological” AND “pain management” AND “transcultural nursing”, combined as MESH term and keyword. The inclusion criteria applied in this study were peer-reviewed articles in English that discussed non-pharmacological pain management and transcultural nursing. Articles published within the last five years (2017-2021). Research studies were carried out in various areas that specifically examined non-pharmacological pain management with transcultural nursing approach. This study was a quantitative study with a randomized controlled trial (RCT) and full text method. The first author performs an initial database search and articles for review. We used the PRISMA Flowchart 2009 (Moher et al., 2010) to record the article review and inclusion process (see Figure 1). An initial search of four databases yielded 1.210 results. After that, we collected all articles and removed duplicated articles. The source was excluded by title and abstract if it was not a peer-reviewed research study or related to non pharmacological pain management with transcultural nursing approach. The next step was to narrow the selection of articles based on the year of publication and the research contexts. After the remaining articles were assessed for significant non pharmachological pain management findings, 7 papers were selected for final inclusion (Figure 1).

**Figure 1.**
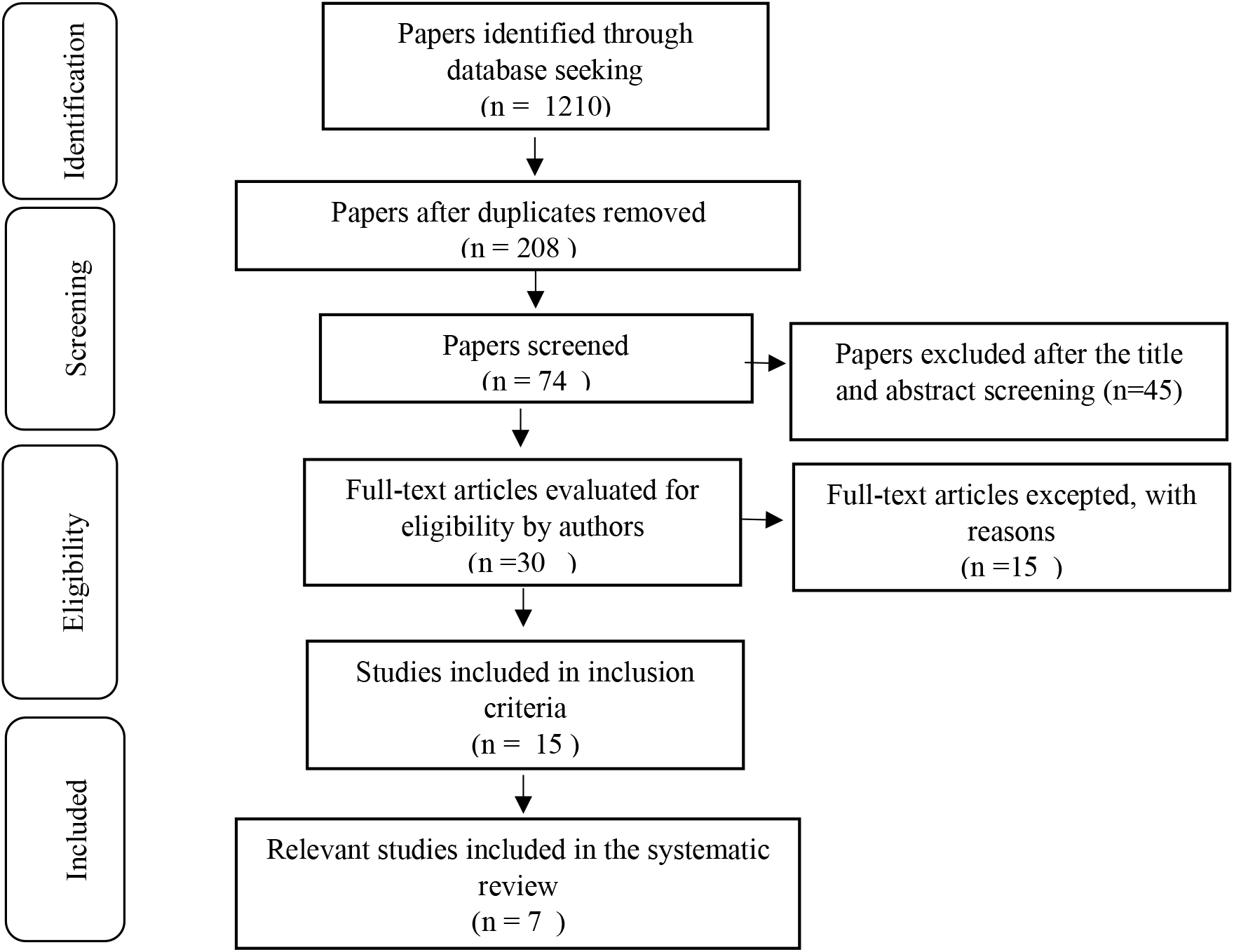
PRISMA Flowchart of Literature Search and Screening Process

## 3. RESULTS AND DISCUSSION

### 3.1. Ontological Studies on Pain

Pain is a protective mechanism of the body to protect and provide red flags about disturbances in the body. Pain mechanism occurs due to the presence of stimuli received by the pain receptors and is altered in the form of impulses that are delivered to the pain centers in the cortex of the brain. After the process of concentrating pain, the impulse is returned to the periphery in the form of pain perception. The stimuli received by the pain receptors can come from various factors such as : mechanical, chemical and thermal (Black & Hawk, 2014). Pain is influenced by biological, psychological, and social factors at varying degrees. Pain is the most common symptom and reason for patients to seek medical care and common problems that need to be addressed.

The research of Andersson et al found that 29% of inpatients reported experiencing moderate to severe pain during rest and 41% reported experiencing moderate to severe pain during activity. The lack of proper pain management causes the pain problem to continue until after discharge from the hospital. Pain can even last more than 3 months and become a chronic pain that can affect the patient’s quality of life, work and sleep. It also causes patients to seek medical care repeatedly, which indirectly leads to increased costs on the Healthcare system (Lin et al., 2021). Therefore, the provision of proper pain management for inpatients is an important issue that cannot be ignored.

Factors affecting patient pain control show a relationship between the patient’s age, gender, disease category, hospitalization day, emotional distress, and the patient’s pain level on the day of discharge. Those factors significantly associated with the patient’s pain on the day of discharge are age, gender, disease category, length of hospitalization, degree of pain on the day of admission, emotional stress on the day of admission, and emotional stress on the day of discharge (P<.001). There is a significant negative correlation between the age and degree of pain of the patient, the higher the age the lower the degree of pain. Degree of pain in men is significantly lower than in women. The degree of pain of surgical patients on the day of discharge is significantly higher than that of gynecological patients and gynecological patients is significantly higher than that of internal medicine patients. While in the hospital, the level of pain on the day of admission, the emotional stress on the day of admission, and the emotional distress on the day of returning home were significantly correlated positively with the level of pain. The longer the patients stay in the hospital, the higher the level of pain on the day of admission, the greater the emotional stress on the day of admission and the greater the emotional stress on the day of exit (Lin et al., 2021).

### 3.2 Epistemological Studies on pain and transcultural nursing

#### 3.2.1 Definition of pain

Pain, according to IASP (*International Association for the Study of pain*) is defined as an unpleasant sensory and emotional experience associated with tissue damage or potentially tissue damage (Ariwibawa et al., 2017). Pain is not only nervous activity, but also a high level of cognitive processes. It is influenced by biological, psychological, and social factors at different levels.

#### 3.2.2 Pain mechanism

Pain is a mixture of physical, emotional and behavioral reactions. The three physiological components in the experience of pain are reception, perception and reaction. The pain-producing stimulus sends impulses through peripheral nerve fibers. The nerve fibers enter the spinal medulla and through one of several nerves and finally come inside the gray mass of the spinal cord. There are pain messages that can interact with inhibitor nerve cells that prevent the pain stimulus from reaching the cerebral cortex, so the brain interprets the quality of pain and processes information about past experiences and knowledge and cultural associations in an effort to perceive pain.

The pain process starting from the stimulation of the nociceptor by the noxious stimulus to the occurrence of the subjective experience of pain is a series of electrical and chemical events that can be grouped into 4 processes, namely transduction, transmission, modulation and perception.

##### 1) Transduction

The process of transduction or activation of receptors is a pain mechanism that starts from the stimulation of the nociceptor by the noxious stimulus in the tissues and will then result in the stimulation of the nociceptor. The noxious stimulus will be transformed into an action potential. Furthermore, the action potential will be transmitted to the neurons of the central nervous system associated with pain.

##### 2) Transmission

Transmission is the first stage of conduction of impulses from the primary afferent neuron to the dorsal cord of the medulla spinalis, in this dorsal cornnu the primary afferent neuron sneezes with the neuron of the central nervous system. Furthermore, the network of neurons will rise upwards in the spinal medulla towards the brainstem and thalamus. Then there is a reciprocal relationship between the thalamus and the higher centers in the brain that regulates the perceptual and affective responses associated with pain and vice versa the perception of pain can occur without nociceptive stimulation.

##### 3) Modulation

The signal modulation process is able to influence the pain process, where the most known signal modulation is in the dorsal cord of the spinal medulla.

##### 4) Perception

Perception is the last process by which pain messages are passed towards the brain and produce unpleasant experiences.

In addition to the four processes of the mechanism of pain, there is a theory of pain control (gate control) proposed by Melzack & Wall (1965 in Potter & Perry, 2006). This theory posits that pain impulses can be regulated or even inhibited by defense mechanisms along the central nervous system. Defense mechanisms can be found in substantive gelatinous cells within the dorsal cord in the spinal medulla, thalamus and limbic systems. This gate control theory explains that pain impulses are delivered when a defense is opened and impulses are inhibited when a defense is closed. Efforts to close those defenses are the basis for pain relief therapy.

A balance of activation of sensory neurons and the descendent control fibers of the brain regulates defense processes. Delta-A and C neurons release substance P to transmit impulses through defense mechanisms. In addition there are mechanoreceptors, beta-A neurons that are thicker and faster to release inhibitory neurotransmitters. If the dominant input comes from delta-A fibers and C fibers, it opens up those defenses and the patient perceives pain. The desert nerve groove releases endogenous opiates, such as endorphins and dinorphins, which are natural pain killers that originate in the body. This neuromodulator closes the defense mechanism by inhibiting the release of substance P. Distraction techniques, counseling and placebo administration are attempts to release endorphins.

#### 3.2.3 Transcultural Nursing

Transcultural nursing is a major direction in nursing that focuses on comparative studies and analyses of different cultures and sub-cultures in the world that value *caring behavior*, nursing services, values, beliefs about healthy illness, as well as patterns of behavior aimed at developing a body of knowledge which is scientific and humanistic to give place to the practice of nursing to a particular culture and a universal culture (Alligood, 2014). This theory of transcultural nursing emphasizes the importance of the role of nursing in understanding client culture. The nurse’s correct understanding of the client’s culture, whether individual, family, group, or community, can prevent culture shock and culture imposition. Cultural shock occurs when outsiders (nurses) try to study or adapt effectively to a certain cultural group (client) while culture imposition is the tendency of health workers (nurses), either secretly willing to blatantly impose cultural values, beliefs, and habits /behaviors that they have and individuals, families, or groups of other cultures because they believe that their culture is higher than that of other groups.

Theory of transcultural nursing sunrise in the name of sunrise this model symbolizes the essence of nursing in transcultural which explains that before providing nursing care to clients (individuals, families, groups, communities, institutions), nurses must first have knowledge of the worldview of dimensions and cultures and social structures.

The role of nurses in transcultural nursing theory is to distinguish between the care system carried out by ordinary people and the professional care system through nursing care. The existence of the role of the nurse is described by Madeleine M. Leininger. Therefore the nurse must be able to make nursing decisions and action plans that will be given to the community. If adjusted to the nursing process, it is the planning stage of nursing actions.

Transcultural Nursing Care Assessment developed by Madeleine M. Leininger explains nursing care in a cultural context depicted in the form of a sunrise (Sunrise Model). The nursing action given must pay attention to the 3 principles of nursing care namely (Alligood, 2014):

#### a. Culture care preservation/maintenance

This stage is done when the patient’s culture does not conflict with health. The planning and implementation of nursing is provided according to the relevant values that the client already has so that the client can improve or maintain his health status, for example the culture of exercising every morning.

#### b. Culture care accommodation/negotiation

This is done to help clients adapt to certain cultures that are more beneficial to health. Nurses help clients to choose and determine other cultures that are more supportive of improving health, for example pregnant clients have abstinence from eating fishy smells, so fish can be replaced with other sources of animal protein.

#### c. Culture care repatterning/restructuring

This stage is carried out when the culture is detrimental to health status. Nurses seek to restructure the lifestyles of clients who normally smoke to not smoke. The pattern of the chosen life plan is usually the one that is more profitable and corresponds to the beliefs adopted.

Conceptual model developed by Madelaine M. Leininger in explaining nursing care in a cultural context is depicted in the form of a sunrise (Sunrise Model). The management of nursing care is carried out from the stage of assessment, nursing diagnosis, planning, implementation and evaluation. Assessment is the process of collecting data to identify a client’s health problems according to the client’s cultural background. The assessment is designed based on seven components in the “Sunrise Model”

a. Technological factors Health technology allows individuals to choose or get offers to solve problems in health care. Nurses need to examine: Perceptions of healthy illness, habits of seeking health care or overcoming health problems, reasons for seeking health help, reasons for clients to choose alternative treatments and client perceptions of the use and use of technology to address these health problems.
b. Religious and philosophical factors of life (religious and philosophical factors) Religion is a symbol that results in a very realistic view for its adherents. Religion provides a very strong motivation to get the truth above all else, even above one’s own life. Religious factors that must be studied by the nurse are: religion adopted, marital status, the client’s perspective on the causes of the disease, the way of treatment and religious habits that have a positive impact on health.
c. Kinship and social factors The nurse at this stage should examine the factors: full name, nickname, age and place of date of birth, gender, status, family type, decision-making in the family and the client’s relationship with the head of the family
d. Cultural values and life ways Culturalvalues are something that is formulated and established by adherents of a culture that is considered good or bad. Cultural norms are rules that have a limited application to adherents of related cultures. The one thing that needs to be studied in this factor is the position and position held by the head of the family, the language used, the eating habits, the food that is abstained in a sick condition, the perception of the sick persons related to daily activities and the habit of cleaning oneself.
e. Political and legal factors The applicable hospital policies and regulations are everything that affects the activities of individuals in cross-cultural nursing care (Andrew and Boyle, 1995). This is what needs to be studied a few others: rules and policies related to visiting hours, the number of family members who can wait, the payment method for clients who are hired.
f. Economic factors Client who was treated at the hospital used the material resources he had to pay for his illness to get better soon. Economic factors that must be studied by the nurse include: client’s work, sources of medical expenses, savings owned by the family, costs from other sources such as insurance, reimbursement from the office or joint ventures between family members
g. Educational factors The client’s educational background is the client’s experience in taking the highest formal path today. The higher the client’s education, the client’s confidence is usually supported by rational scientific evidence and the individual can learn to adapt to a culture that suits his or her health condition. The things that need to be studied at this stage are: the level of education of the client, the type of education as well as his ability to actively learn independently about the experience at least so that it does not repeat itself. The assessment of the seven components can be seen in figure 2 below :

**Figure 2.**
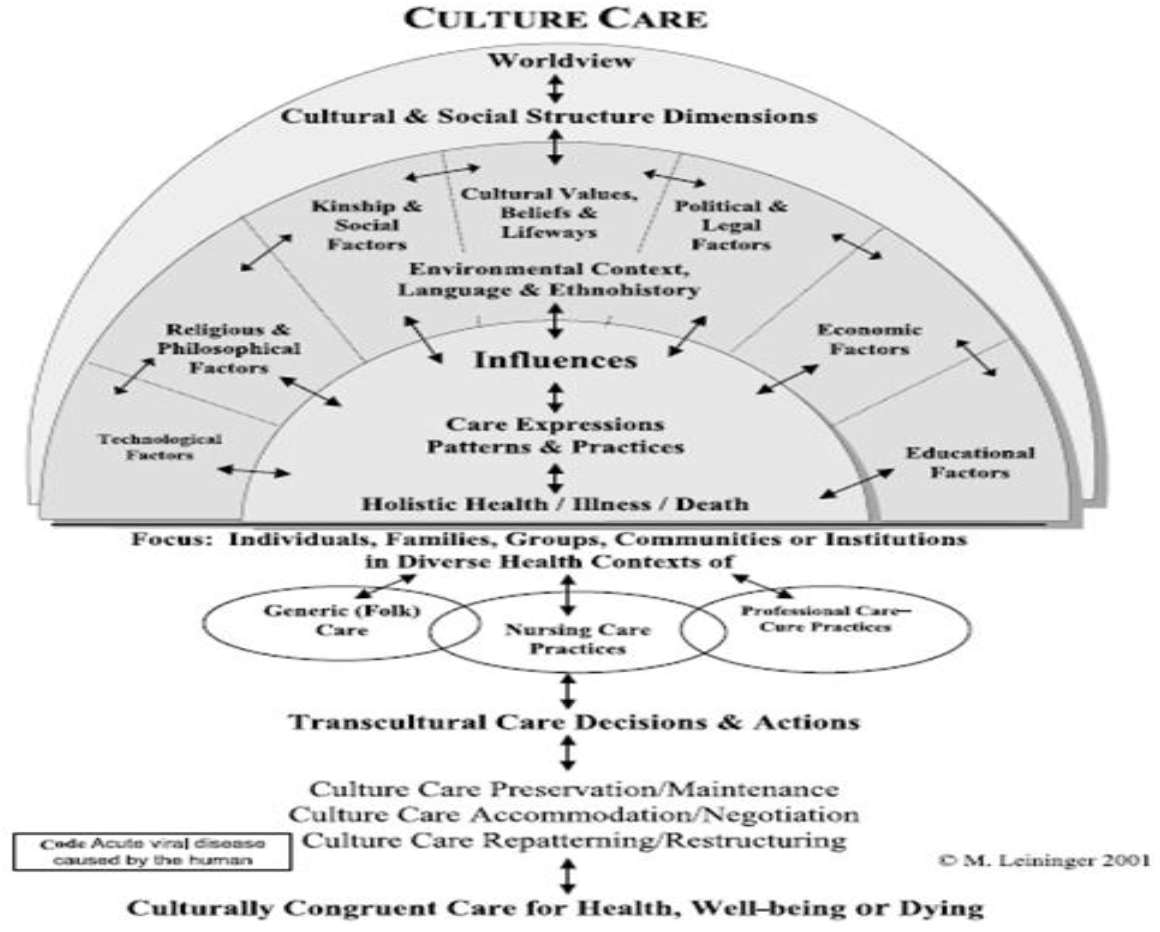
Sunrise Model

### 3.3 Philosophy of non-pharmacological pain management intervention based on axiological perspective

Based on the results of several literature reviews in table 1 of nonpharmacological pain management that can be done to reduce pain, including: cold application (Seweid et al., 2021), heat therapy (Kwon et al., 2022), positioning, acupuncture and reflexiology (Kia et al., 2021), slow deep breathing relaxation exercise (Jarrah et al., 2022), distraction (Ibitoye et al., 2019), Tai Chi (R. Wang et al., 2022) dan music therapy (C. Wang & Tian, 2021).

**Table 1.** The results of the review of articles based on the Axiological perspective approach (n=7)

| No | Title, Author, Year | Method<br>(Design, Subject, Variable, Instrument, Analyze/DSVIA) | Result |
| --- | --- | --- | --- |
| 1 | Effect of cold application on incisional pain associated with incentive spirometry after coronary artery bypass graft surgery (Seweid et al., 2021) | **D** : A crossover study design<br>**S** : A convenience sample of 60 adults, fully conscious postoperative CABG patients newly admitted to open heart surgery ICU were included in the current study. Patients were included if they met the following criteria: First planned CABG surgery and having the ability to self-report and determine the quality of pain.<br>**V** : cold application and incisional pain associated with incentive spirometry after coronary artery bypass graft surgery<br>**I** : subjective pain assessment sheet (Pain intensity scale, Pain distress scale), Objective pain assessment sheet (Physiological indicators of pain assessment, Critical Care Pain Observation Tool (CPOT)<br>**A** : Univariate binary logistic regression | Cold application reduces incisional pain associated with incentive spirometry in patients undergoing coronary artery bypass grafting |
| 2 | Effects of Heating Therapy on Pain, Anxiety, Physiologic Measures, and Satisfaction in Patients Undergoing Cystoscopy (Kwon et al., 2022) | **D** : a single-blinded, single-center, randomized controlled trial (RCT)<br>**S** : patients who were to undergo cystoscopy for the diagnosis and treatment. The inclusion criteria were as follows: (1) patients older than 18 years; (2) had | Heating therapy reduced both subjective and objective pain and anxiety in the experimental group compared to the control group. Heating therapy also decreased the systolic and diastolic blood |
|  |  | sufficient intellectual capacity to communicate and comprehend the details of the study; (3) understood the purpose of the study; (4) provided informed consent; and (5) had urinary problems such as blood in the urine, urinary tract infections, overactive bladder, pelvic pain, etc. We excluded patients diagnosed with bladder cancer as their diagnoses may induce anxiety. Patients who were already in pain were excluded because their pain could not be differentiated from pain due to cystoscopy.<br>V : Heating Therapy on Pain, Anxiety, Physiologic Measures, and satisfaction<br>I : a combined numeric rating scale and face rating Scale, the State-Trait Anxiety Inventory (STAI), electronic OMRON M3 Comfort® HEM e7134-E BP Monitor, the Korean version of the Client Satisfaction Questionnaire (CSQ-8 Korean)<br>A : Mann Whitney U test, analysis of covariance (ANCOVA) | pressure and pulse rate in the experimental group compared to the control group. Women reported significantly greater satisfaction than men. |
| 3 | Nurses' use of non pharmacological pain management methods in intensive care units: A descriptive cross-sectional study (Kia et al., 2021) | D : descriptive cross-sectional design<br>S : 224 nurses who working in 16 ICU wards (i.e., 14 general ICU wards and 2 cardiac surgery and burn ICU wards) across ten university-affiliated hospitals in Northern Iran<br>V : Demographic variables, non pharmacological pain management methods.<br>I : Clinical and demographic questions, | A moderate number of ICU nurses used non-pharmacological pain management methods (55.8 %). The most common method used by nurses was repositioning (M = 2.72), while methods such as acupuncture and reflexology were used less frequently. Furthermore, the most common obstacles to the |
|  |  | The non-pharmacological pain management checklist , a researcher-developed checklist.<br>A : Variance and *t*-test | use of non-pharmacological pain management methods were nurses' fatigue (M = 2.92) and multiple responsibilities (M = 2.91). Demographic variables such as age, gender, educational level, and work experience were not significantly associated with the use of non-pharmacological pain management methods. |
| 4 | The effect of slow deep breathing relaxation exercise on pain levels during and post chest tube removal after coronary artery bypass graft surgery<br>(Jarrah et al., 2022) | D : quasi-experimental study<br>S : fifty post-CABG patients were conveniently selected from a cardiac intensive care unit. he patients were randomly assigned to either an intervention group or a control group. A total of 25 patients were assigned into the experimental group who received slow deep breathing relaxation Exercise (SDBRE) alongside the conventional care before CTR. The remaining 25 patients constituted the control group (50%) that had CTR following conventional care<br>V : slow deep breathing relaxation exercise on pain levels<br>I : Visual Analogue Scale (VAS)<br>A : Bivariate analyses of chi-squared test of independence, Fisher exact test, and independent groups t-test, Non-parametric Mann-Whitney U, Kruskal-Wallis | Using SDBRE during CTR is an effective technique for reducing pain which can minimize the need for analgesics and their associated adverse effects. |
| 5 | The use of distraction as a pain management technique among nurses in a North-central city in Nigeria.<br>(Ibitoye et al., 2019) | D : A cross-sectional and descriptive design<br>S : The study participants were selected using purposive sampling method. They are all registered nurses/registered midwives working in 7 separate wards in the hospital. These wards include the male and female medical and surgical wards, pediatric, stroke and psychiatric wards. sample size was calculated as 163 nurses/midwife.<br>V : distraction and pain management technique<br>I : The questionnaires<br>A : The chi-square test | Majority of the study participants were familiar with distraction and its use in pain management (98.8%). The majority of the participants (97.5%) indicated they use distraction as a pain management technique, and they (61.3%) believed that distraction can be effective, without administering any pharmacological agent. The nurses (84%) mostly used distraction to manage post-operative pain. |
| 6 | Effect of Tai Chi Quan on the Pressure Pain Thresholds of Lower Back Muscles in Healthy Women<br>(R. Wang et al., 2022) | D : randomized controlled trial<br>S : Sixty healthy women aged 18–40 years were randomly assigned to Tai Chi group or control group. The Tai Chi group received 40-minute practice, and the control group received 5-minute sham Tai Chi Quan practice and 35-minute rest.<br>V : Tai Chi Quan and Pressure Pain Thresholds of Lower Back Muscles<br>I : A hand-held digital manometer, pain catastrophizing scale (PCS)<br>A : paired sample *t*-test, Mann-Whitney | The pressure pain thresholds of test points in the Tai Chi group showed substantial improvements after exercise, whereas those in the control group did not improve. Overall, the pressure pain thresholds in the Tai Chi group significantly increased compared with the control group (longissimus thoracis: $p = 0.000$, iliocostalis lumborum: $p = 0.000$, multifidus muscle: $p = 0.000$, quadratus lumborum: $p = 0.012$, gluteus medius: $p = 0.000$ and supraspinous ligament: $p = 0.000$) |
| 7 | Music Intervention to Orthopedic Patients: A Possible Alternative Solution to Control Pain | D : The experimental design<br>S : 38 postoperative orthopedic patients were equipped with pocket-size MP3 players with prerecorded music tracks | It was found that during the intervention of music, the pain was significantly reduced from 5.40 to 2.98. There was a slight relationship between listening time and pain |

| No | Title, Author,<br>Year | Method<br>(Design, Subject,<br>Variable, Instrument,<br>Analyze/DSVIA) | Result |
| --- | --- | --- | --- |
|  | (C. Wang & Tian, 2021) | (instrumental and lyrical) in Hindi, English, and Urdu.<br>V : Music Intervention, pain<br>I : Instruments and Music Choice<br>A : descriptive statistics (frequency and percentage) and paired sample *t*-tests. | relief. It was also found that the feedback was extremely positive and each patient suggested the use of music to others with 96.6% recommendation. |

Based on axiological studies, almost all nonpharmacological therapies in pain management can reduce pain intensity. In addition, some of these nonpharmacological pain management can reduce anxiety and psychological response (Wu et al., 2017), improve sleep quality (Bagheri et al., 2021).

## 4. CONCLUSION

Pain is an unpleasant sensory and emotional experience associated with tissue damage both actual and potential. Pain is the subjective experience of the patient. Race, culture and ethnicity are important factors in a person responding to pain. It is necessary to manage nonpharmacological pain by considering culturally sensitive care. Some non-pharmacological nursing interventions to reduce pain include: cold application, heat therapy, positioning, acupuncture and reflexology, slow deep breathing relaxation exercise, distraction, Tai Chi, music therapy, and Benson Relaxation Technique.

## Data Availability

All data produced in the present work are contained in the manuscript

## ACKNOWLEDGEMENT

The authors thank to the Faculty of Nursing Universitas Airlangga for the facilities in this study

